# Forecastability of infectious disease time series: are some seasons and pathogens intrinsically more difficult to forecast?

**DOI:** 10.1101/2025.04.29.25326677

**Authors:** Lauren A. White, Tomás M. León

**Affiliations:** California Department of Public Health, Center for Infectious Diseases, Division of Communicable Disease Control

**Keywords:** forecastability, predictability, Shannon entropy, spectral density, infectious disease modeling, forecasting, COVID-19, influenza

## Abstract

For infectious disease forecasting challenges, individual model performance typically varies across space and time. This phenomenon raises the question: are there properties of the target time series that contribute to a particular season, location, or disease being more difficult to forecast? Here we characterize a time series’ future predictability using a forecastability metric that calculates the spectral density of the time series. Forecastability of syndromic influenza hospital admissions for the state of California varied widely across seasons and was positively correlated with peak burden. Next, using archived U.S. state and national forecasts targeting laboratory-confirmed COVID-19 and influenza hospital admissions, we investigated the relationship between forecastability and: (i) population size of the forecasting target, and (ii) forecast performance as measured by mean absolute error, weighted interval score (WIS), and scaled relative WIS. Forecastability increased with increasing population size of the forecasting target, and forecasting performance generally improved with higher forecastability when controlling for population size across scales. These preliminary results support the idea that some targets and respiratory virus seasons may be inherently more difficult to forecast and could help explain inter-seasonal variation in model performance.

**Author summary:** Could intrinsic properties of an epidemiological time series help explain why a particular season, location, or disease is more difficult to predict in the future? To answer this question, this analysis uses a measure of a time series’ future predictability called “forecastability,” which describes the inherent uncertainty or surprise in the signal. Influenza and COVID-19 hospital admissions had higher forecastability scores for locations with larger population sizes, possibly due to larger counts leading to smoother time series. At the same time, forecasting performance generally improved for time series with higher forecastability scores when controlling for population size, suggesting that this metric is helpful for understanding ease of forecasting. These preliminary results support the idea that some epidemiological targets and respiratory virus seasons may be inherently more difficult to forecast and could help explain why forecasting model performance changes across different respiratory virus seasons.

## Introduction

There have been numerous collaborative forecasting efforts across multiple years and diseases including influenza, COVID-19, dengue, and Ebola (1–4). These forecasting challenges or “hubs” provide unified submission targets for modeling teams, evaluate the performance of individual model predictions, and synthesize predictions from multiple submitting teams into a unified, ensemble model. Historically, individual models fail to outperform such ensembles consistently through time (2,5). Forecast performance also tends to decline during periods of rapid change, and certain forecasting targets have been more difficult to forecast than others, e.g., COVID-19 cases compared to COVID-19 deaths (5,6). However, it remains unclear which constituent factors of a model contribute to higher performance, nor is it uncommon for individual model performance to vary within and across seasons and locations (5). Here we ask the inverse question about the underlying forecasting target itself: is there something intrinsic about the qualities of infectious disease time series that make them more or less challenging to forecast?

In the infectious disease forecasting literature, quantile forecasts are commonly evaluated with scoring metrics like mean absolute error (MAE) and weighted interval score (WIS). MAE describes the absolute difference between the forecasted value and the observed value, while WIS also accounts for the dispersion of the forecast’s prediction interval. Both of these metrics scale with the absolute size of their target (7). This naturally gives more weight to locations with a higher disease burden. In contrast, applying a log-transformation prior to evaluation means that forecasts are scored based on relative error instead of absolute error, giving equal weight to location targets with smaller population sizes (8). Model performance is commonly evaluated against a baseline model which propagates incidence forward based on values from the prior time point with uncertainty based on changes in incidence observed in prior time intervals (2).

“Forecastability” describes the future predictability—or the inherent uncertainty or surprise—in the signal of a stationary time series. Other disciplines have used various measures of entropy to characterize the complexity of time series and their propensity for future predictability (9–12). Scarpino and Petri proposed permutation entropy as a way of measuring the predictability limitations of infectious disease time series (13). Predictability decreased with increasing time series length, illustrating that more time series history did not necessarily correlate with increased predictability according to that specific metric. This study also found that different diseases had unique slopes of predictability vs. time series length, suggesting that permutation entropy was responding to specific signatures in each disease time series. Mills et al. 2025 calculated permutation entropy of a rolling window of COVID-19 weekly incident cases and compared with time-varying forecast performance to understand how predictability affects forecast value for decision making (14). In the context of decision making, the ensemble model generally had greater relative performance especially at longer forecast horizons, even during periods of lower predictability.

Another proposed metric for measuring forecastability calculates the Shannon entropy of the spectral density of the time series (15). In the realm of forecasting evaluation, the 2020 West Nile Virus Forecast Challenge used Shannon entropy to assess uncertainty in the ensemble model forecast of binned case counts (16). This metric is invariant to shifting or scaling, with values near 0% corresponding to white noise and values near 100% corresponding to a single sinusoidal curve (Figure 1). For example, S&P 500 returns which have minimal autocorrelation may have values near 1%, while mean temperature time series with significant autocorrelation lags expected at six and twelve months might have a forecastability closer to 35% (15). This metric does not interact with forecast specifications like model type, forecast horizon, or loss function—depending only on the target time series itself (15). Although there are potentially several ways to define forecastability, we will hereafter use the term “forecastability” to refer to the definition outlined in Goerg (2013).

**Figure 1.**
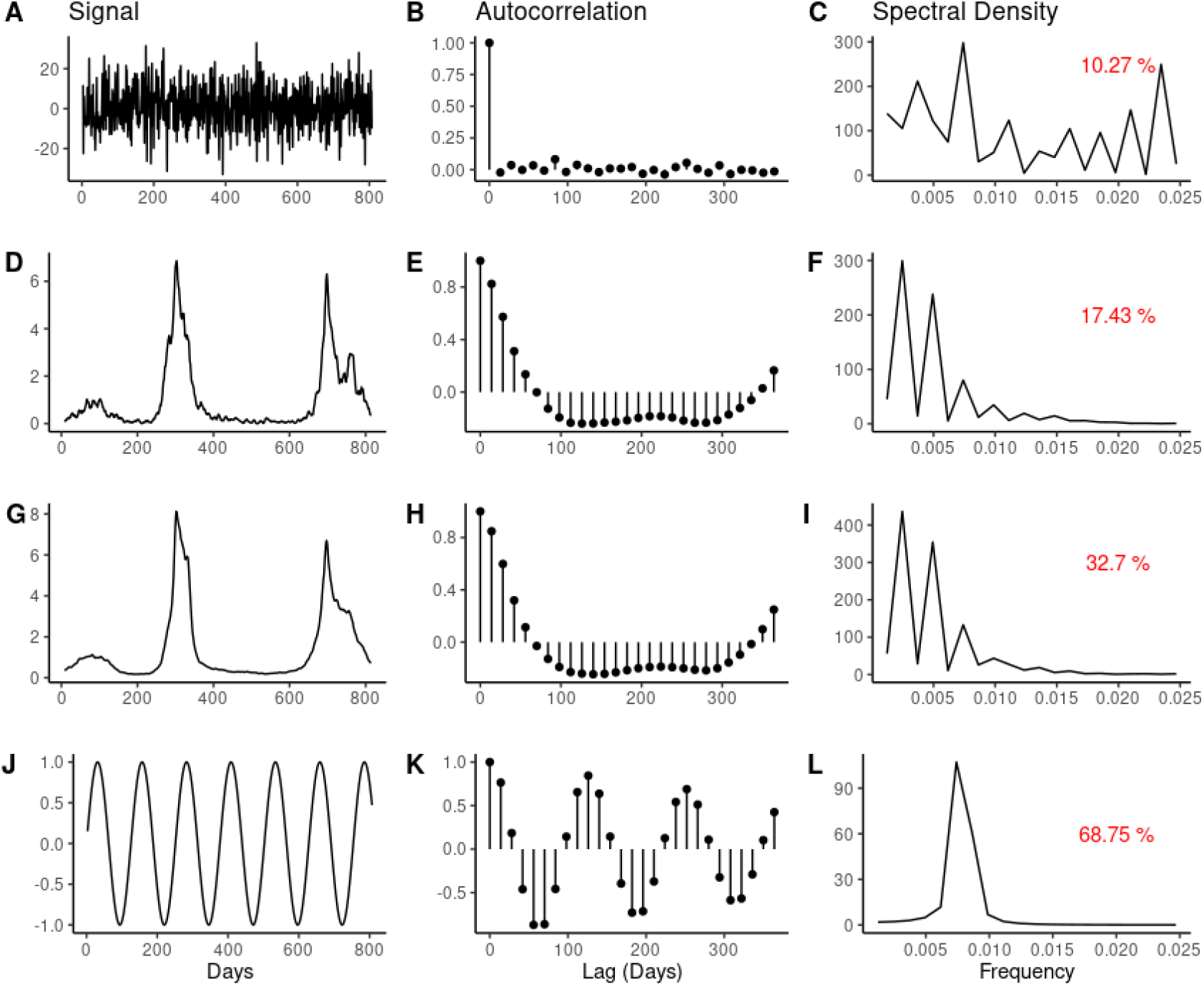
Correspondence between example time series (first column), their autocorrelation values (second column) and spectral density in the frequency domain (third column). The red text in the third column corresponds to a forecastability score of the original signal. Time series increase in their “forecastability” from top to bottom, ranging from purely random (first row) to perfectly sinusoidal (fourth row). The second row presents weekly influenza admissions per 100K for the state of Maryland and the third row corresponds to weekly influenza admissions per 100K for the entire United States. The x-axis of the signal column represents days since mandatory reporting for influenza hospital admissions was required beginning February 2, 2021. The autocorrelation and spectral densities were calculated for the entire time series displayed in the first column, and for ease of visual display, autocorrelation lags at intervals of 14 days were plotted with a maximum lag of 365 days displayed. For consistency, all sample time series are differenced once prior to calculating the forecastability score.

Other studies have found that forecasting performance improves for larger jurisdictions –e.g., states vs. counties (6) and larger vs. smaller counties (17). We further hypothesized that: (1) as population size increases, forecastability of the time series should also increase; and (2) as forecastability increases, the WIS of the natural log transformed forecast should decrease (i.e., forecast performance should improve with higher forecastability, when controlling for population size). We explored the relationship between population size of the representative time series and forecastability: (1) at a county and state-level for California for syndromic influenza hospital admissions data from the California Department of Healthcare Access and Information (HCAI); (2) at a U.S. state and national level for laboratory-confirmed hospital admissions data for COVID-19 and influenza derived from Health and Human Services (HHS) Patient Impact and Hospital Capacity Data System/National Hospital Safety Network (NHSN) (18). With the latter, we investigated the relationship between forecastability and forecast performance at the state and national scales using archived forecasts from the COVID-19 Forecast Hub (https://covid19forecasthub.org/) and FluSight Forecast challenge (https://github.com/cdcepi/FluSight-forecast-hub)—two collaborative forecasting initiatives that have targeted laboratory-confirmed hospital admissions for COVID-19 and influenza respectively (5,19). We believe this is the first application linking this forecastability metric to infectious disease forecast performance across diseases and seasons.

## Methods

### Data

California county and state level syndromic influenza hospitalization data were derived from HCAI for the 2000-2022 respiratory virus seasons. State and national, laboratory-confirmed, COVID-19 and influenza hospitalization admission data were obtained from HHS/NHSN via Delphi COVIDcast for the 2022-2024 respiratory virus seasons (18,20). Seasons were classified by the standard influenza season definition of MMWR week 40 to week 39—specifically, October 2, 2022-September 30, 2023 for the 2022-2023 season and October 1, 2023-April 30, 2024 for the 2023-2024 season, where only voluntary reporting continued after April 30, 2024. To assess sensitivity of results to the season timing, an alternative season definition from July 1, 2022-July 30, 2023 & July 1, 2023-April 30, 2024 was also explored.

State and national population size estimates were taken from the 2021 US Census Bureau estimates (21). California county population estimates were taken from 2020 California Department of Finance estimates (22).

To identify the predominant influenza subtype for historical California seasons, the most frequently sequenced subtype was taken from historic Respiratory Laboratory Network (RLN) and clinical sentinel laboratory surveillance data (23).

Historical forecasts for state and national COVID-19 and influenza targets were obtained from the COVID-19 Forecast and the FluSight Forecast challenge, respectively, for the 2022-2023 and 2023-2024 respiratory virus seasons (5,19). Specifically, we focused on a comparison of baseline and ensemble models, as well a few select models that were submitted consistently to both forecasting hubs across both seasons (e.g., CMU-TimeSeries, PSI-DICE, SGroup-RandomForest, UMass-trends_ensemble). Ensemble models from the modeling hubs combine all available models for a given target date and location and usually provide the most robust performance (2,5). In contrast, baseline models offer a reference point for model performance by propagating historical values and uncertainty forward (2).

### Forecastability

Time series can be analyzed in the frequency domain through spectral analysis to reveal the constituent frequencies of a signal. This is reflected by the relationship between the autocovariance function and the spectral density, which are reciprocal Fourier pairs of one another. The autocovariance function of a stationary timeseries *x*_*t*_ is given by:

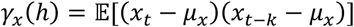

Where 𝔼 is the expectation operator, *h* is the lag value that takes on any integer value, and μ_*x*_ = 𝔼 [*x*_*t*_] or the mean of *x*_*t*_. The spectral density for the same process, *x*_*t*_, for a given frequency, ω, is achieved by taking the Fourier transform of the autocovariance γ_*x*_:

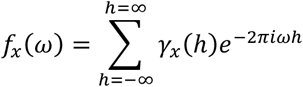

Where 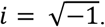.

Shannon entropy measures the inherent uncertainty of a probability distribution for a discrete random variable *X* for set χ:

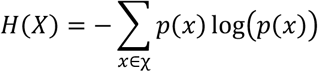

where *p*(*x*) is the probability of state *x* being observed in set χ. For a continuous random variable, Shannon entropy is approximated by differential entropy:

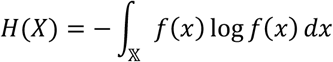

for a continuous random variable *X* with probability density function *f*(*x*) and support 𝕏 ∈ ℝ. Combining the concepts of spectral density and differential entropy, the forecastability of a stationary timeseries *x*_*t*_ is more formally defined as:

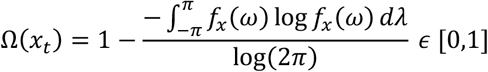

where *fx* (ω) is the normalized spectral density of *x*_*t*_ or 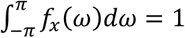 (15). Here log(2π) represents the expected differential entropy value of a white noise signal, and so Ω(*x*_*t*_) would equal zero if and only if *x*_*t*_ corresponded to true white noise.

Time series were assessed for stationarity with both the Kwiatkowski-Phillips-Schmidt-Shin (KPSS) and the Augmented Dickey-Fuller (ADF) tests at an alpha = 0.01. For comparability across geographies and seasons, time series were differenced once before calculating the forecastability score. The metric of forecastability was calculated using the Omega function of the foreCA package (15).

### Forecast evaluation

Forecasts and their corresponding truth data were first transformed via the natural logarithm to control for the differing magnitude in burden across target locations (8). Following Bosse et al. (2023) and general convention, we added small positive quantity (*a* =1) to the truth and predictions data prior to scoring, to account for potential values of zero while maintaining a monotonic transformation (8). Point forecast error was assessed with mean absolute error (MAE), which takes the absolute difference between a model’s median point predictions ŷ _*i=*1:*N*_ and the set of observed outcomes *y*_*i=*1:*N*_ for a given model, location, and season such that 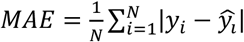

Probabilistic forecast accuracy was assessed with the weighted interval score (WIS), which is a metric that compiles the performance of a quantile or interval-based forecasts over a range of prediction intervals (7). The single, interval score for a forecast *F* with outcome *y* for prediction interval (1 − α) × 100% is given by:

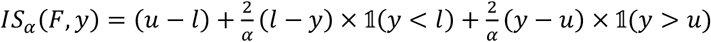

Where 𝟙 is the indicator function, *u* is the upper prediction limit and *l* is the lower prediction limit. The score resolves into three components: *u* − *l* represents a penalty incurred for width or dispersion of the prediction interval, 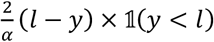 conveys a penalty for underprediction, and 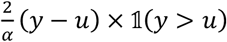 conveys a penalty for overprediction.

The WIS then sums intervals scores prediction intervals at different levels (1 − α_1_) < (1 − α_2_) < ⋯ < (1 − α_*K*_) such that:

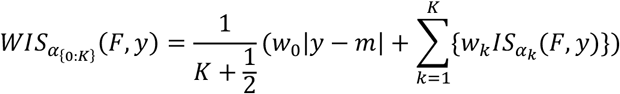

Where *K* is the total number of quantiles included in the WIS, *m* is the predictive median forecast value, and *w*_*k*_ is a non-negative, unnormalized weight, often set to: 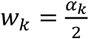 and 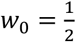 (7).

In this case, both MAE and WIS values approximate relative error since these were computed on the natural logarithm of the forecasts and their respective targets (8).

Lastly, models were scored using relative skill, θ_*i*_, which computes model rankings by taking the geometric mean of all possible pairwise comparisons across all models, as given by:

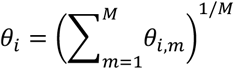

where θ_*i*,*m*_ represents the mean score ratio of model *i* and model *m* out of all possible models *M*. Because this metric is based on pairwise comparisons, it only produces a score when models contribute to the same target for the same date(s) and can therefore help to control for forecast missingness (24). Here we present scaled relative WIS results which normalizes model performance relative to a baseline model; relative WIS values below one indicate that the given model performs better than the baseline model, whereas value greater than one suggest a model performs worse than the baseline model on average. MAE, mean WIS, and scaled relative risk were calculated for each location, season, and model combination across all four week horizons using the scoringutils package in R (24). All analyses were completed in R (25).

### Ethics Statement

The California Health and Human Services Agency Committee for the Protection of Human Subjects (CPHS) has determined that this research (project number 2024-210) is classified as exempt under the federal Common Rule. This decision is issued under the California Health and Human Services Agency’s Federal wide Assurance #00000681 with the Office of Human Research Protections (OHRP).

## Results

When looking at California county and state-level syndromic influenza hospitalizations, for which more seasons of data were available, some respiratory virus seasons were more “forecastable” than others, with the greatest differences occurring for locations with larger population sizes (Fig 2A). For example, forecastability ranged from 19.5% to 41.6% for the state of California for the 2021-2022 and 2017-2018 seasons respectively. Across seasons from 2000-2022, there was a positive relationship between forecastability and both cumulative burden and peak burden of each respective season with an adjusted R-squared of 0.56 and 0.81 respectively (Fig 2B, Supplemental Fig 1). The season with highest peak weakly admissions, cumulative burden, and forecastability was an H3N2 season (Fig 2B, Supplemental Fig 1). Two other H3N2 majority seasons (i.e., 2003-2004, 2005-2006) were outliers with higher-than-expected forecastability given their peak burden. The two seasons bridging the 2009 swine influenza pandemic (2008-2010) were outliers with lower-than-expected forecastability given their burden.

**Figure 2.**
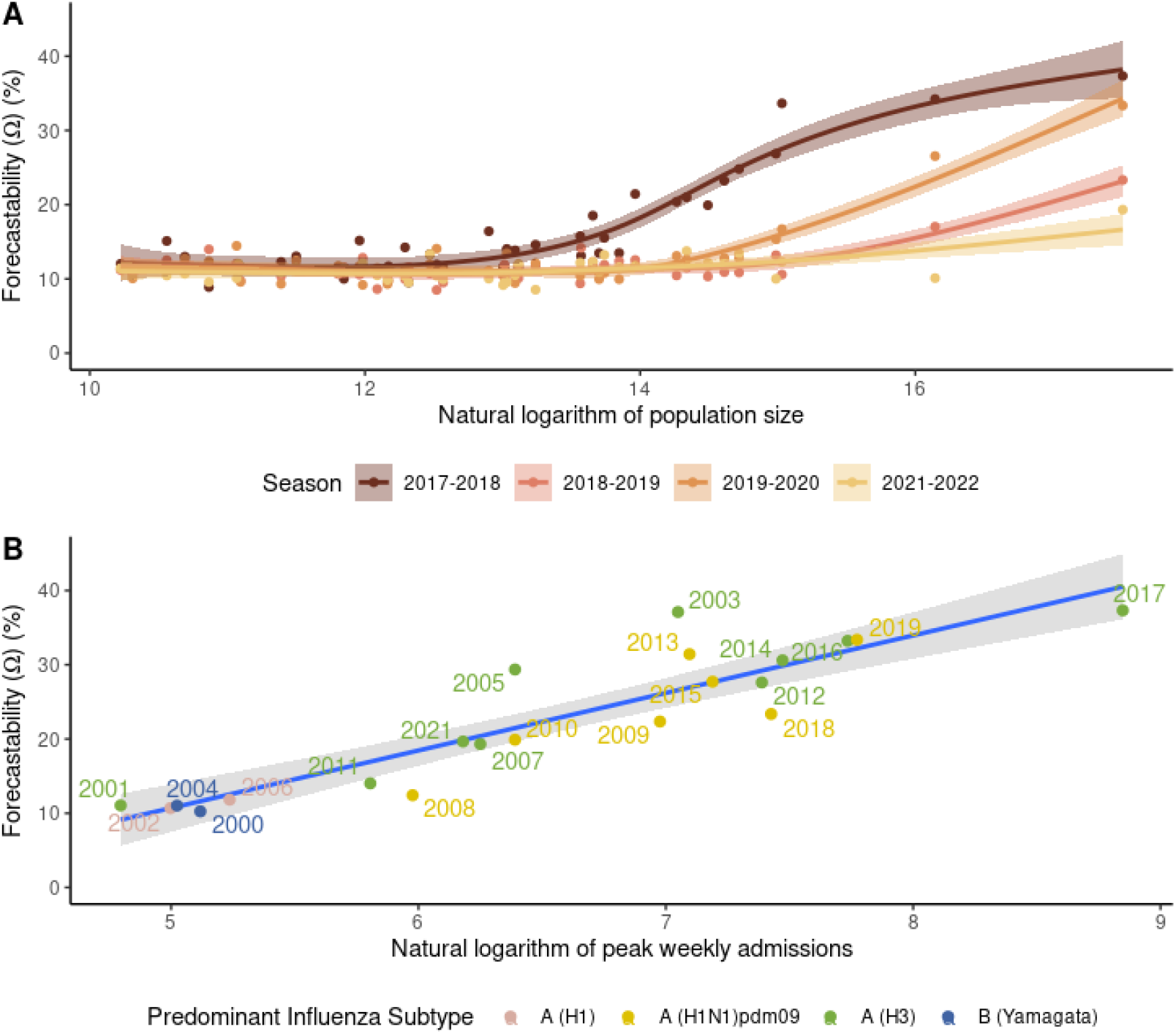
Examples of variation in forecastability by season for syndromic HCAI influenza hospitalizations. **(A)** The relationship between forecastability and population size for the state of California and constituent counties by respiratory virus season, derived from HCAI data from 2017-2022 (2020-2021 excluded because of the COVID-19 pandemic) and fitted with a generalized additive model (GAM) for visualization; **(B)** forecastability vs. the natural log of peak influenza weekly admissions for the state of California derived from syndromic HCAI data from 2001-2022 and fitted with a linear regression (*β*= 7.49, p < 0.001). The year labels in the plot correspond to the first year of the MMWR season, e.g., 2001 corresponds to the 2001-2002 respiratory virus season.

For U.S. state and national laboratory confirmed admissions, forecastability of the time series increased with target population size for both COVID-19 and influenza across U.S. states and nationally (Fig 3A). This pattern was generally of the same order of magnitude for both COVID-19 and influenza, and similar across both the 2022-2023 & 2023-2024 seasons (Fig 3A). As seen in prior studies (6,17), forecast performance generally improved for more populous targets, i.e., for natural log transformed forecasts, MAE and WIS on the log scale decreased with increasing population size for the ensembles (Fig 3B-D) and most individual models (Supplemental Fig 2). Except for the 2022-2023 influenza season, MAE and WIS for the ensemble on the log scale also decreased with higher forecastability, i.e., forecast performance improved with higher forecastability (Fig 3B &C, Table 1). For example, for COVID-19 in 2022-2023 season, forecastability values ranged from 10% (Idaho) to 30% (United States) with corresponding decreases in relative error of MAE from 25% to 21% and relative error of WIS from 26% to 14% for the ensemble. In contrast, baseline model performance showed no such relationship with forecastability, with non-significant or positive slope estimates (Fig 3B & C, Table 1).

**Table 1.**
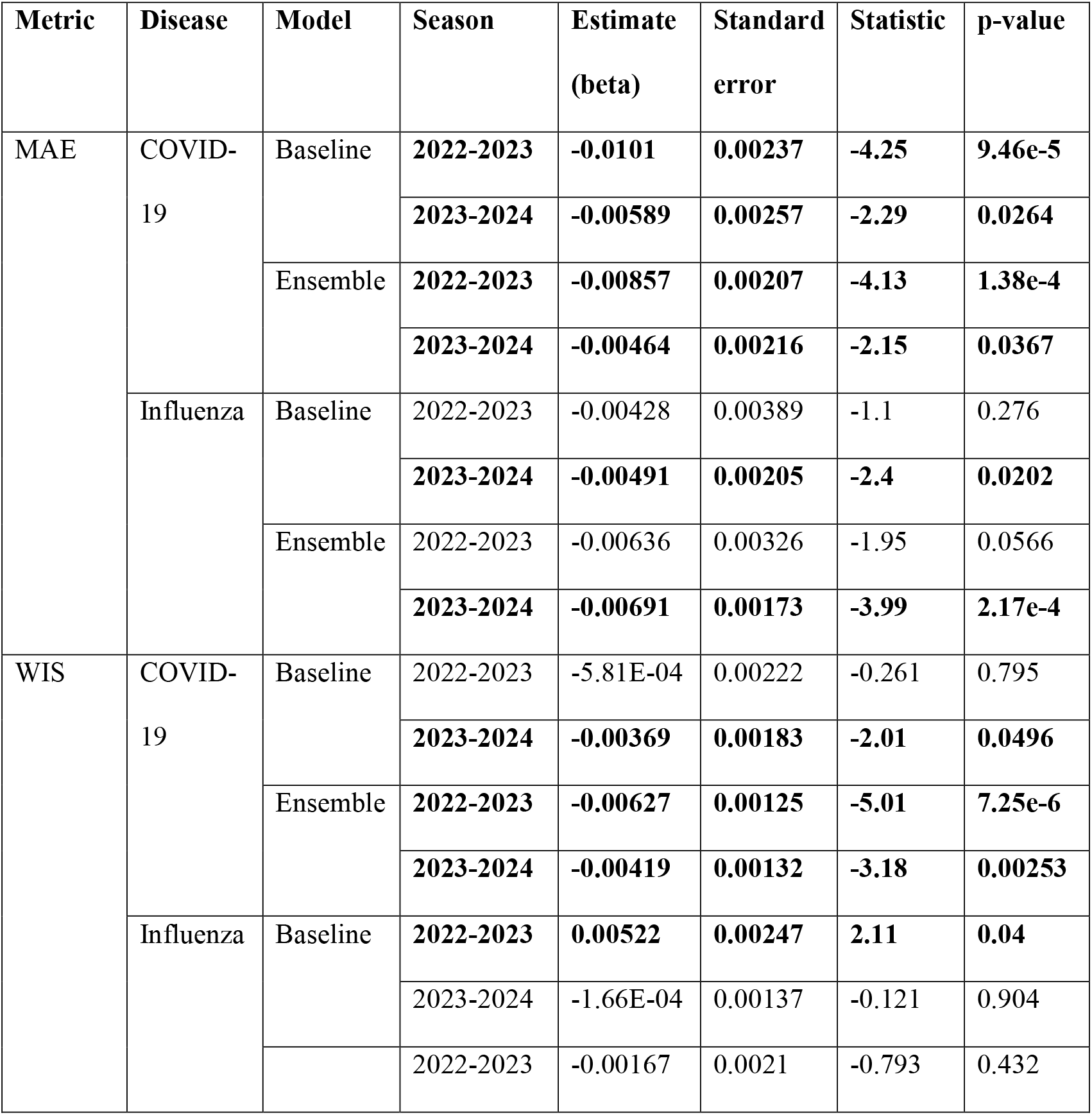
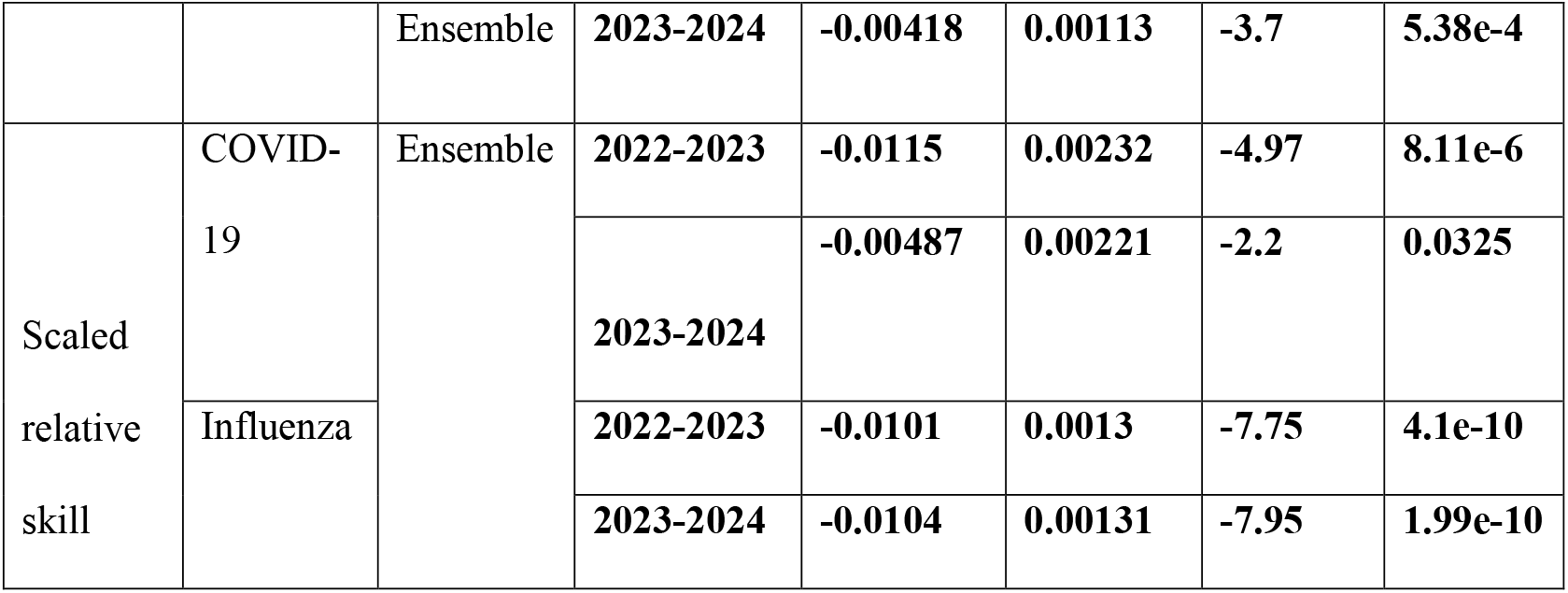
Slope estimates from linear model fits for forecastability (Ω) vs. forecast performance for the ensemble and baseline models targeting laboratory-confirmed (HHS/NHSN) COVID-19 and influenza admissions at the U.S. state and national scales as shown in Figure 2. Values are shown with three significant figures. Bolded rows are those statistically significant at a p = 0.05 threshold.

**Figure 3.**
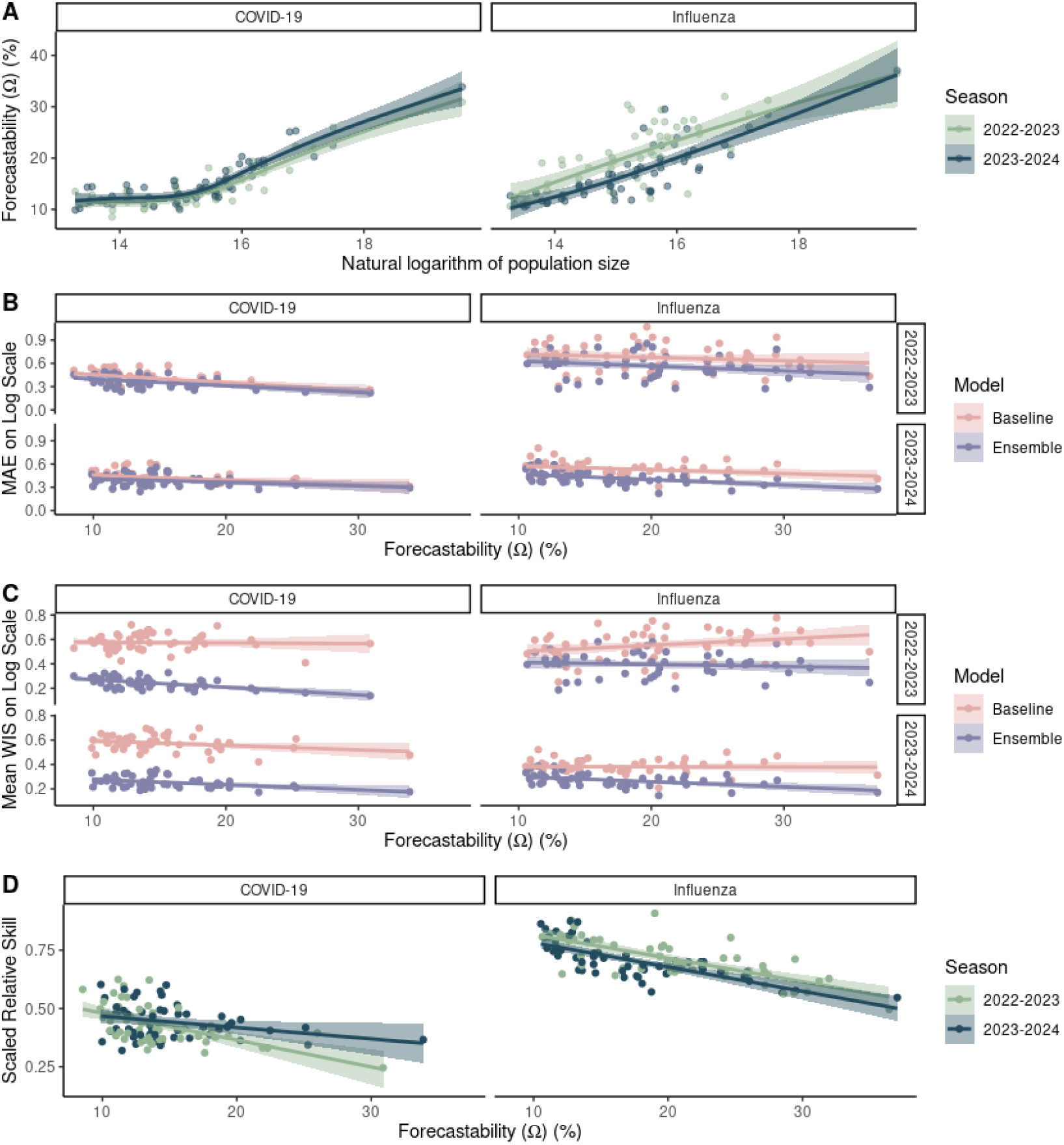
The relationship between population size, forecastability, and forecast performance across two respiratory virus seasons for laboratory-confirmed (HHS/NHSN) COVID-19 and influenza admissions at the U.S. state and national scales. **(A)** Forecastability (Ω) vs. the natural logarithm of population size. Forecast performance for the baseline and ensemble models as measured by: **(B)** MAE and **(C)** mean WIS and **(D)** scaled relative skill for the ensemble model.

Across both COVID-19 and influenza ensembles, rates of change of MAE per unit forecastability ranged from -0.0046 to -0.0086, i.e., every one percent unit increase in forecastability yielded a ∼0.46-0.86% decrease in MAE relative error (Table 1). Rates of change of WIS per unit forecastability ranged from -0.0016 to -0.0062 for a corresponding decrease in WIS relative error of ∼0.13-0.62% per unit forecastability (Table 1). Relationships between the ensembles’ scaled relative skill and forecastability were significant for both seasons and both diseases (Fig 3C, Table 1) with increases in relative skill ranging from -0.0048 to -0.011 per unit of forecastability. Overall, these findings were robust to alternative definitions of seasonal timing (Supplemental Figure 3, Supplemental Table 1).

For lab-confirmed COVID-19 and influenza hospital admissions, the metric of forecastability was very susceptible to data reporting frequency (Fig 4) and the amount of data smoothing (Fig 5). When compared to a time series of rolling weekly admissions with a daily reporting frequency, time series down sampled to weekly reporting frequency had a decreased forecastability score. While this observed effect increased with population size of the measured time series, population size explained less of the variance in forecastability for the weekly vs. the daily sampled time series (Fig 4). The relationship between forecastability and the amount of data smoothing exhibited asymptotic dynamics—beginning with lower forecastability scores for daily reporting that improved to a plateau as the length of the rolling sum window increased (Fig 5).

**Figure 4.**
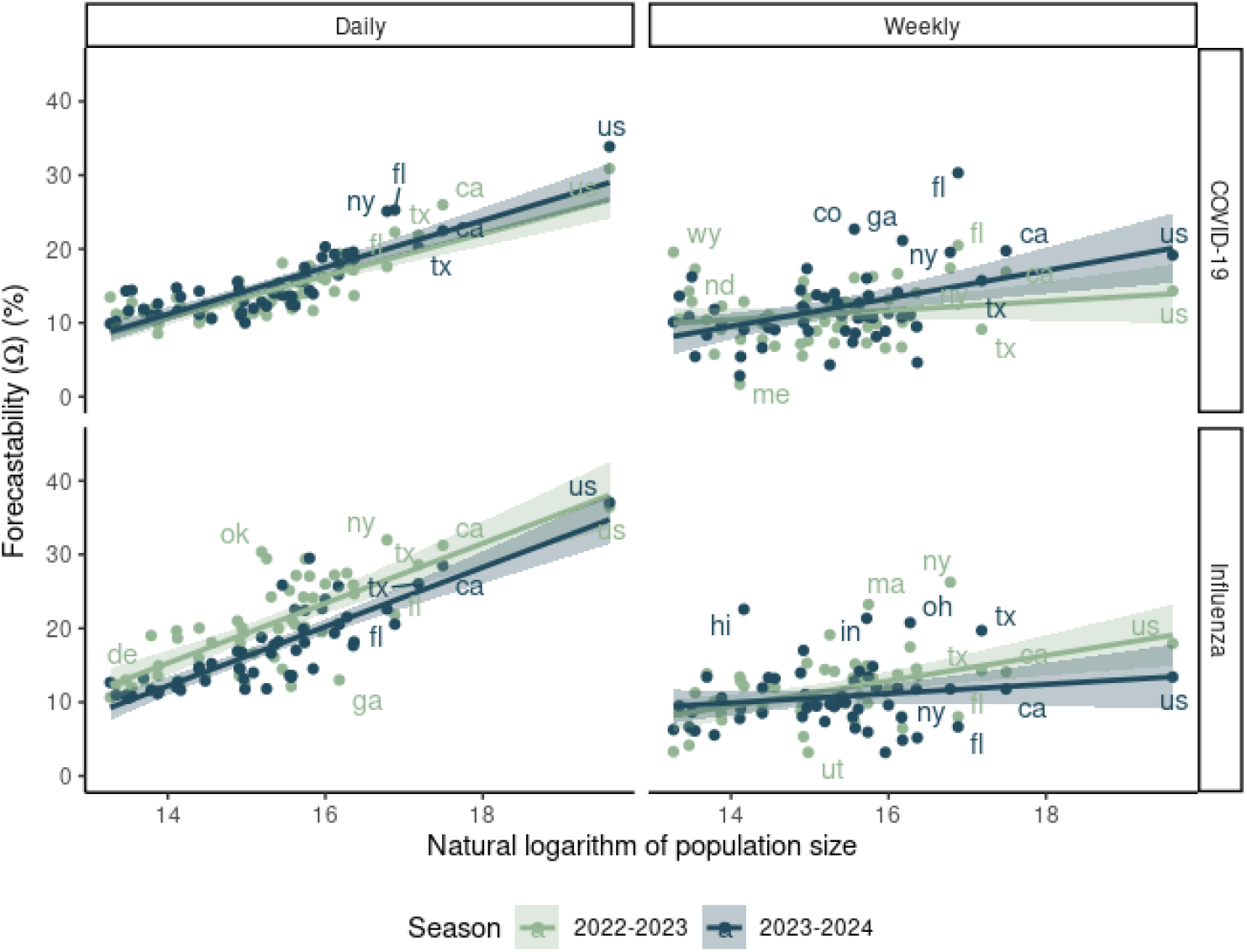
Comparison of forecastability scores vs. natural logarithm of population size for daily vs. weekly reporting (columns) for COVID-19 vs. influenza laboratory-confirmed hospital admissions (rows) from state and national HHS/NHSN data for the 2022-2023 and 2023-2024 seasons.

**Figure 5.**
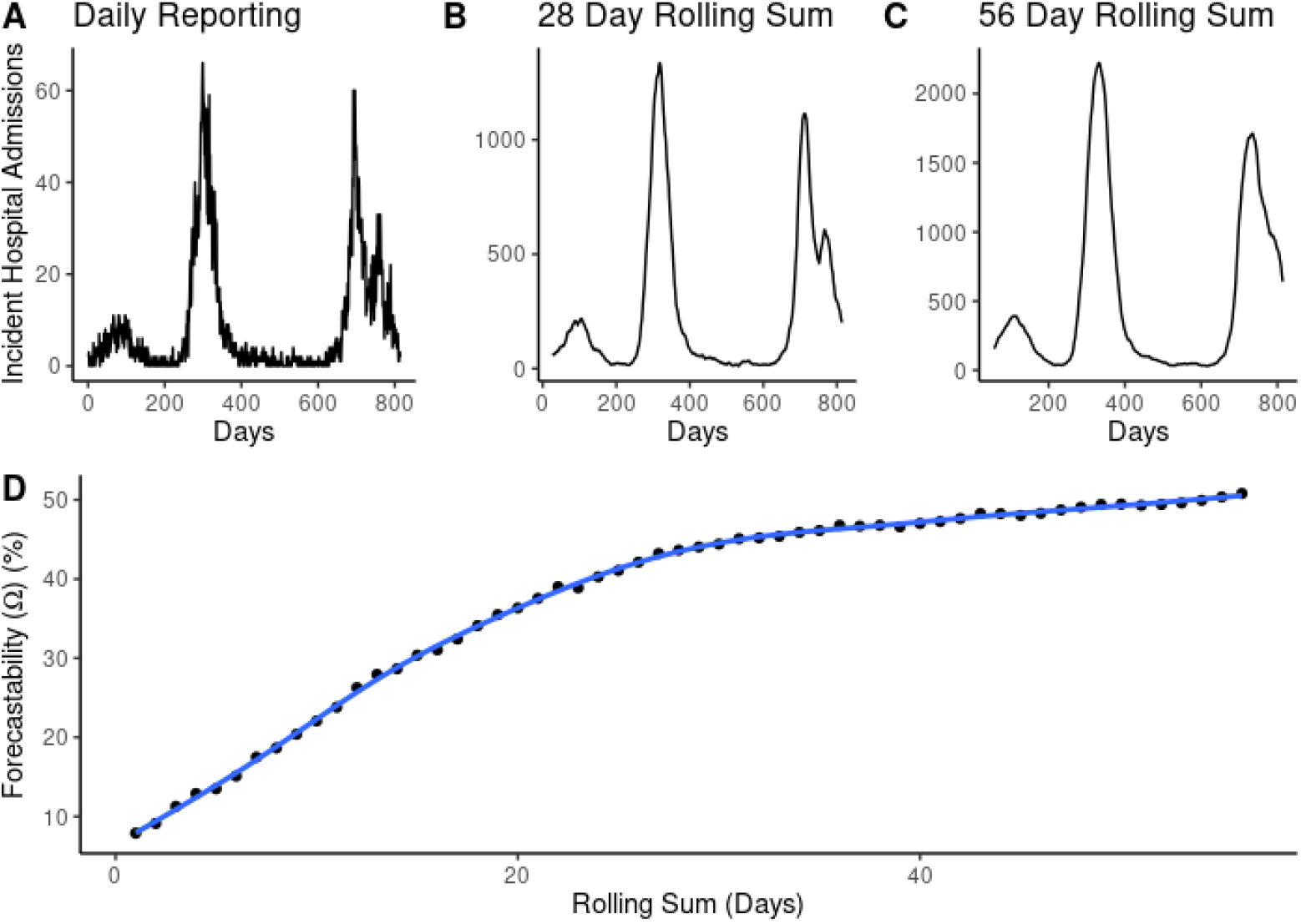
Exploration of the relationship between the amount of signal smoothing and forecastability score. (A-C) Time series of incident influenza hospital admissions with daily reporting vs. 28-day or 56-day rolling sums for the state of Maryland. The x-axis of the signal column represents days since mandatory reporting for influenza hospital admissions was required beginning February 2, 2021. (D) Forecastability scores calculated on incident influenza hospitalization time series aggregated by the days of aggregation designated in the x-axis. The trend line is a Generalized Additive Model (GAM) fit on forecastability as a function of the rolling sum window (days) with a cubic spline basis function.

## Discussion

Using a simple signal processing metric to characterize the predictability of infectious disease time series, we showed that there are differences in the measured forecastability of time series across diseases (i.e., COVID-19 vs. influenza, Fig 3A) and across respiratory virus seasons for a given disease (Fig 2). Moreover, these observed differences in measured forecastability correlated with resulting forecast performance. Our results echoed prior work that found a relationship between population size of the forecast target and forecast performance (Fig 3B & C, Supplementary Fig 2) (6,17), but our work further suggests that forecastability increases with population size (Fig 2A, Fig 3A), and forecast performance improves for time series with higher forecastability scores when controlling for population size (Fig 3D). Notably, scaled relative skill of the ensemble against the baseline model improved with increasing forecastability of the target time series across all four combinations of season and disease (Fig 3D), and these gains came from improvements in ensemble performance rather than changes in the baseline model performance, which remained relatively invariant to the time series’ forecastability (Fig 3C, Table 1). This observed relationship between forecastability and population size reflects general intuition, i.e., that larger populations should have signals closer to the mean field, with corresponding weaker high frequency components, resulting in a higher forecastability score.

Interestingly, there appears to be a strong relationship between peak seasonal burden and forecastability (Fig 2B), which may point to seasons with more singular, sharper peaks having higher forecastability per this particular metric. However, this finding contrasts with the general pattern that forecasts struggle most during periods of rapid change, such as during the growth phase or at the epidemic peak (5,6). Although anecdotal, patterns in forecastability across seasons matches what might be expected based on epidemiological understanding: forecastability was lower than expected for disrupted influenza seasons (e.g., during the 2009 swine influenza pandemic and in the wake of the COVID-19 pandemic) (Fig 2B). This also follows intuition, as one would expect that periods of lower overall burden would exhibit more epidemic stochasticity, while peak periods would better approximate mean field behavior. This observed variability in forecastability across seasons could help explain why there has been such variability in both model rankings across years and the ability (or inability) of models able to outperform the baseline model for any given season (5).

The patterns of forecastability observed here raise a couple of questions about the practical implementation of forecasting challenges for infectious diseases. The first is that forecastability is strongly affected by data frequency (e.g. daily vs. weekly) and amount of data smoothing (e.g., daily vs. weekly rolling sum). Although the increase in forecastability observed with population size is less evident with lower frequency data (Fig 4) and forecastability has a non-linear relationship with the amount of data smoothing (Fig 5), we were not able to tie these observation directly to forecast performance. Daily hospital admissions data have been publicly available at certain times for COVID-19 and influenza (and were briefly a formal target for COVID-19), but we do not know which models were training on daily vs. weekly data. This may matter practically since there are often changes in infectious disease data continuity through time in terms of targets, reporting frequency, and reporting definitions (e.g., influenza shifting from influenza like illness (ILI) to confirmed hospitalizations during the COVID-19 pandemic, changes in NHSN reporting requirements from daily to weekly in November of 2024). It also highlights that forecasting a completely new signal (e.g., FluSight 2022-2023) remains challenging regardless of the features of the time series.

The second notable pattern is that there is a consistent plateau of forecastability at about 10% for smaller geographies, regardless of the disease, and that certain seasons were more or less predictable according to the forecastability metric (Figure 2A, 3A). Since the baseline model showed no significant relationship with forecastability (Fig 3C, D), this raises the question of whether forecasting challenges should also be: (1) controlling for some version of signal predictability based on geographic scale or (2) controlling for season predictability post-hoc when scoring forecasts. The latter might matter more for comparing model performance across seasons than within a season. Finally, although there has been increased interest in providing local level forecasts in the wake of the COVID-19 pandemic (17), this also raises questions about infectious disease surveillance for smaller geographies and the inherent feasibility of providing real time infectious disease modeling intelligence at these scales.

Forecastability is an appealing metric because it simplifies a complex time series into a single value; however, the interpretation of this dimension reduction remains challenging. Since forecastability is measured in the frequency domain, it is sometimes difficult to understand which qualities of a time series directly translate to a more forecastable signal. Another limitation of this analysis is that we measured the forecastability of an entire season, while forecast metrics presented here represent mean scores of numerous individual forecasts through time—each occurring at time points with imperfect/incomplete information for the season up to that given date. This metric only reflects the underlying infectious disease time series itself, which ignores the other potential covariates that could form the complete information set of a forecast (e.g., the value of knowing hospitalizations when forecasting deaths) (14). This analysis also does not address potential underlying differences in clinical reporting patterns or biases across geographies and for different diseases. For example, providers might be less likely to test for and diagnose influenza during off season months, or with increased usage of multiplex testing, influenza detections may increase incidentally as a result.

There are numerous other ways to measure or categorize the properties of time series and future work could explore these metrics in an infectious disease context (11,12). Other methods that consider more local behavior of the time series at the time of the forecast (e.g., shapelet analysis, method of analogues) might also address this limitation of a global time series metric being applied to a summation of local forecast scores (26,27). For example, although COVID-19 and influenza had roughly similar relationship and scale between population size and forecastability (Fig 3A), COVID-19 had noticeably lower relative WIS across both seasons compared to influenza (Fig 3D)—suggesting that this metric is not capturing all features relevant to forecasting performance. This work could contribute to improved selection of baseline models for scoring forecasts (e.g., (28)), better cross-seasonal comparisons of forecasting hub results, and an increased understanding of how forecasts contribute to decision making at different thresholds of uncertainty and predictability (14). We hope this analysis sparks further research into delineating the qualities of infectious disease time series themselves—or the underlying “information set” of forecasts—that may help further explain forecasting performance across seasons and diseases.

## Data availability

Code and underlying data to reproduce the analyses are available at: https://github.com/cdphmodeling/forecastability.

Laboratory-confirmed hospital admissions data are publicly available data sets through HHS/NHSN for state: https://healthdata.gov/Hospital/COVID-19-Reported-Patient-Impact-and-Hospital-Capa/g62h-syeh/about_data and facility-level time series: https://healthdata.gov/Hospital/COVID-19-Reported-Patient-Impact-and-Hospital-Capa/anag-cw7u/about_data

Syndromic influenza hospitalization data derived from the California Department of Healthcare Access and Information (HCAI) contain individual level patient data and so are only available upon a data request: https://hcai.ca.gov/data/request-data/

## Authors contributions

L.W.: conceptualization, formal analysis, methodology, validation, visualization, writing— original draft, writing—review and editing; T.L.: conceptualization, data curation, methodology, supervision, writing—review and editing.

## Funding

This work was supported by the California Department of Public Health. The findings and conclusions in this article are those of the author(s) and do not necessarily represent the views or opinions of the California Department of Public Health or the California Health and Human Services Agency.

This study used the California Patient Discharge Dataset. The interpretation and reporting of these data are the sole responsibility of the authors. The authors acknowledge the California Department of Healthcare Access and Information for compilation of these data.

This work was funded by Centers for Disease Control and Prevention, Epidemiology and Laboratory Capacity for Infectious Diseases, Cooperative Agreement Number 6 NU50CK000539.

## Acknowledgements

The authors thank members of the CDPH Modeling and Advanced Analytics team including Héctor Sánchez Castellanos, Mugdha Thakur, Natalie Linton, Phoebe Lu, and Brent Siegel for conversations and insights that improved these analyses.

## References

1. Biggerstaff M, Alper D, Dredze M, Fox S, Fung ICH, Hickmann KS, et al. Results from the Centers for Disease Control and Prevention’s Predict the 2013-2014 Influenza Season Challenge. BMC Infect Dis. 2016 Jul 22;16:357.

2. Cramer EY, Ray EL, Lopez VK, Bracher J, Brennen A, Castro Rivadeneira AJ, et al. Evaluation of individual and ensemble probabilistic forecasts of COVID-19 mortality in the United States. Proc Natl Acad Sci U S A. 2022 Apr 12;119(15):e2113561119.

3. Johansson MA, Apfeldorf KM, Dobson S, Devita J, Buczak AL, Baugher B, et al. An open challenge to advance probabilistic forecasting for dengue epidemics. Proc Natl Acad Sci U S A. 2019 Nov 26;116(48):24268–74.

4. Viboud C, Sun K, Gaffey R, Ajelli M, Fumanelli L, Merler S, et al. The RAPIDD ebola forecasting challenge: Synthesis and lessons learnt. Epidemics. 2018 Mar;22:13–21.

5. Mathis SM, Webber AE, León TM, Murray EL, Sun M, White LA, et al. Evaluation of FluSight influenza forecasting in the 2021-22 and 2022-23 seasons with a new target laboratory-confirmed influenza hospitalizations [Internet]. medRxiv; 2023 [cited 2024 Jan 30]. p. 2023.12.08.23299726. Available from: https://www.medrxiv.org/content/10.1101/2023.12.08.23299726v1

6. Lopez VK, Cramer EY, Pagano R, Drake JM, O’Dea EB, Adee M, et al. Challenges of COVID-19 case forecasting in the US, 2020–2021. PLOS Comput Biol. 2024 May 6;20(5):e1011200.

7. Bracher J, Ray EL, Gneiting T, Reich NG. Evaluating epidemic forecasts in an interval format. Pitzer VE, editor. PLOS Comput Biol. 2021 Feb 12;17(2):e1008618.

8. Bosse NI, Abbott S, Cori A, van Leeuwen E, Bracher J, Funk S. Scoring epidemiological forecasts on transformed scales. PLOS Comput Biol. 2023 Aug 29;19(8):e1011393.

9. Ponce-Flores M, Frausto-Solís J, Santamaría-Bonfil G, Pérez-Ortega J, González-Barbosa JJ. Time Series Complexities and Their Relationship to Forecasting Performance. Entropy. 2020 Jan;22(1):89.

10. Garland J, James R, Bradley E. Model-free quantification of time-series predictability.

11. Kang Y, Hyndman RJ, Smith-Miles K. Visualising Forecasting Algorithm Performance using Time Series Instance Spaces. 2016;

12. Papacharalampous G, Tyralis H, Pechlivanidis IG, Grimaldi S, Volpi E. Massive feature extraction for explaining and foretelling hydroclimatic time series forecastability at the global scale. Geosci Front. 2022 May 1;13(3):101349.

13. Scarpino SV, Petri G. On the predictability of infectious disease outbreaks. Nat Commun. 2019 Feb 22;10(1):898.

14. Mills C, Irons NJ, Tsui JLH, Sparrow S, Carvalho LM, Kucharski AJ, et al. From metric to action: An evaluation framework to translate infectious disease forecasts into policy decisions [Internet]. medRxiv; 2025 [cited 2025 Aug 7]. p. 2025.07.20.25331802. Available from: https://www.medrxiv.org/content/10.1101/2025.07.20.25331802v1

15. Goerg GM. Forecastable Component Analysis. In: Proceedings of the 30 th International Conference on Machine Learning. Atlanta, Georgia; 2013.

16. Holcomb KM, Mathis S, Staples JE, Fischer M, Barker CM, Beard CB, et al. Evaluation of an open forecasting challenge to assess skill of West Nile virus neuroinvasive disease prediction. Parasit Vectors. 2023 Jan 12;16(1):11.

17. White LA, McCorvie R, Crow D, Jain S, León TM. Assessing the accuracy of California county level COVID-19 hospitalization forecasts to inform public policy decision making. BMC Public Health. 2023 Apr 28;23(1):782.

18. U.S. Department of Health & Human Services. COVID-19 Reported Patient Impact and Hospital Capacity by State Timeseries (RAW) [Internet]. 2024. Available from: https://healthdata.gov/Hospital/COVID-19-Reported-Patient-Impact-and-Hospital-Capa/g62h-syeh/

19. Cramer EY, Huang Y, Wang Y, Ray EL, Cornell M. The United States COVID-19 Forecast Hub dataset. Sci Data. 2022;9(1):462.

20. Reinhart A, Brooks L, Jahja M, Rumack A, Tang J, Agrawal S, et al. An open repository of real-time COVID-19 indicators. Proc Natl Acad Sci. 2021 Dec 21;118(51):e2111452118.

21. U.S. Census Bureau. Population Estimate, July 1, 2021 [Internet]. [cited 2024 Dec 9]. Available from: https://api.census.gov/data/2021/pep/population/variables.html

22. State of California Department of Finance. California 2020 Population by Age [Internet]. [cited 2024 Dec 9]. Available from: https://dof.ca.gov/forecasting/demographics/

23. California Department of Public Health (CDPH) Influenza Surveillance Program. Historic Influenza and Other Respiratory Disease Surveillance Reports [Internet]. [cited 2025 Feb 7]. Available from: https://www.cdph.ca.gov/Programs/CID/DCDC/pages/immunization/flu-reports.aspx

24. Bosse NI, Gruson H, Cori A, Leeuwen E van, Funk S, Abbott S. Evaluating Forecasts with scoringutils in R. arXiv [Internet]. 2022; Available from: https://arxiv.org/abs/2205.07090

25. R Core Team. R: A language and environment for statistical computing [Internet]. Vienna, Austria: R Foundation for Statistical Computing; 2021. Available from: https://www.R-project.org/

26. Srivastava A. DTW+S: Shape-based Comparison of Time-series with Ordered Local Trend [Internet]. arXiv; 2023 [cited 2024 Nov 1]. Available from: http://arxiv.org/abs/2309.03579

27. Viboud C, Boëlle PY, Carrat F, Valleron AJ, Flahault A. Prediction of the Spread of Influenza Epidemics by the Method of Analogues. Am J Epidemiol. 2003 Nov 15;158(10):996–1006.

28. Stapper M, Funk S. Mind the Baseline: The Hidden Impact of Reference Model Selection on Forecast Assessment [Internet]. 2025 [cited 2025 Aug 26]. Available from: http://medrxiv.org/lookup/doi/10.1101/2025.08.01.25332807

