## Supplementary Materials for "Forecastability of infectious disease time series: are some seasons and pathogens intrinsically more difficult to forecast?"

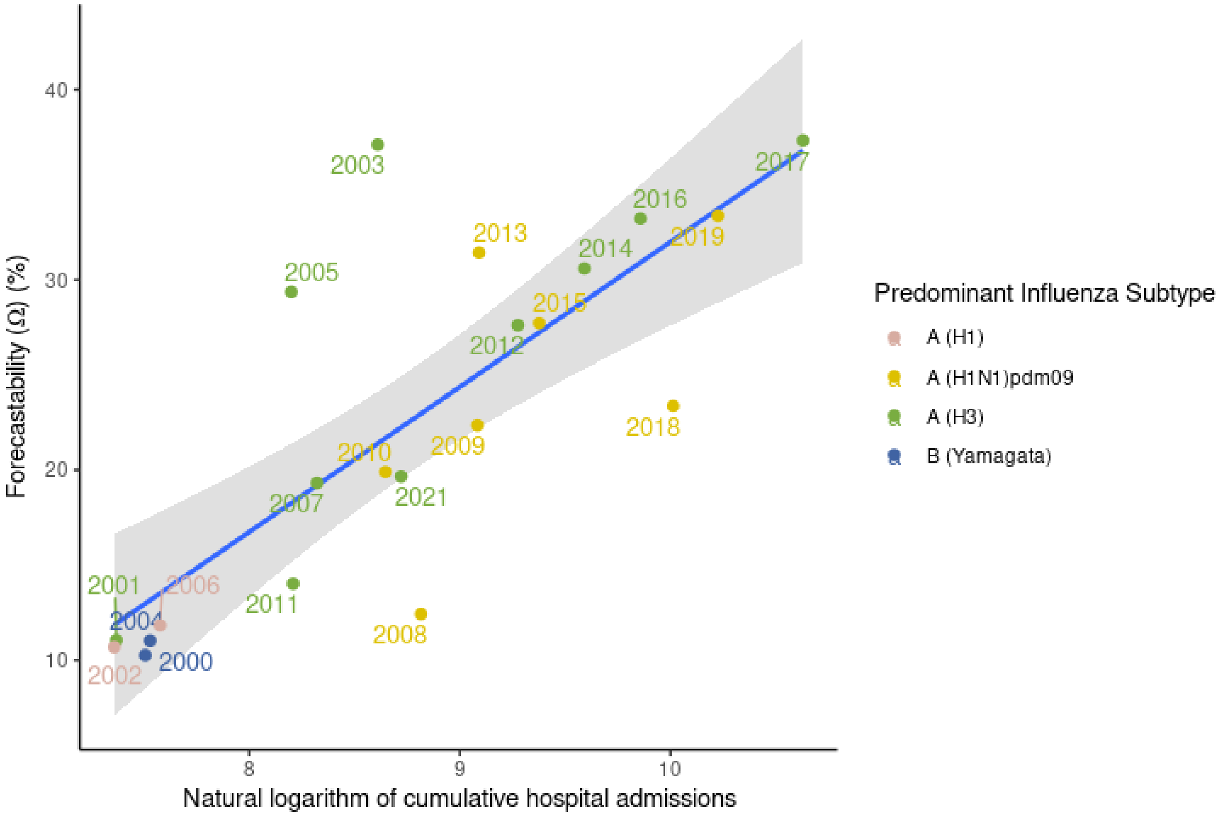


**Supplemental Figure 1.** Forecastability vs. the natural log of cumulative syndromic (HCAI) influenza hospital admissions for the state of California fit with a linear regression ($\beta$= 7.33, $p < 0.001$). Colors of the points correspond to predominant influenza sub-type as captured by influenza surveillance reports from the California Department of Public Health (CDPH) Influenza Surveillance Program.

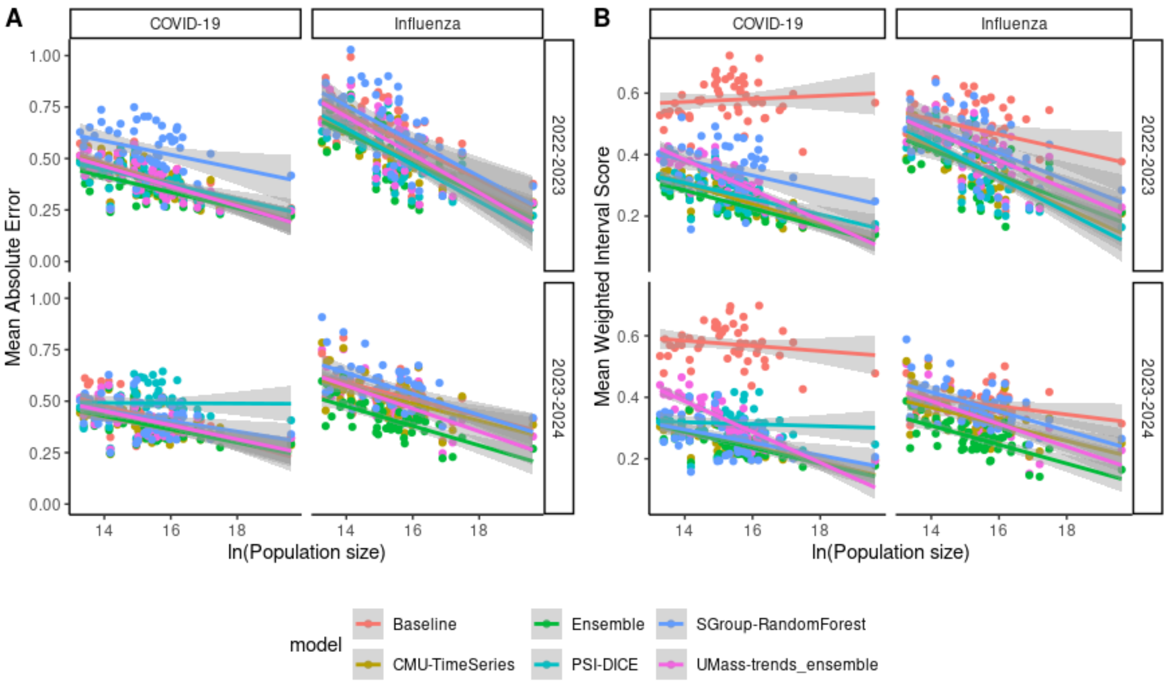


**Supplemental Figure 2.** Target population size vs. forecast performance: **(A)** MAE and **(B)** mean WIS on the log scale for individual models contributing across multiple seasons for the FluSight Challenge and COVID Forecast Hub.

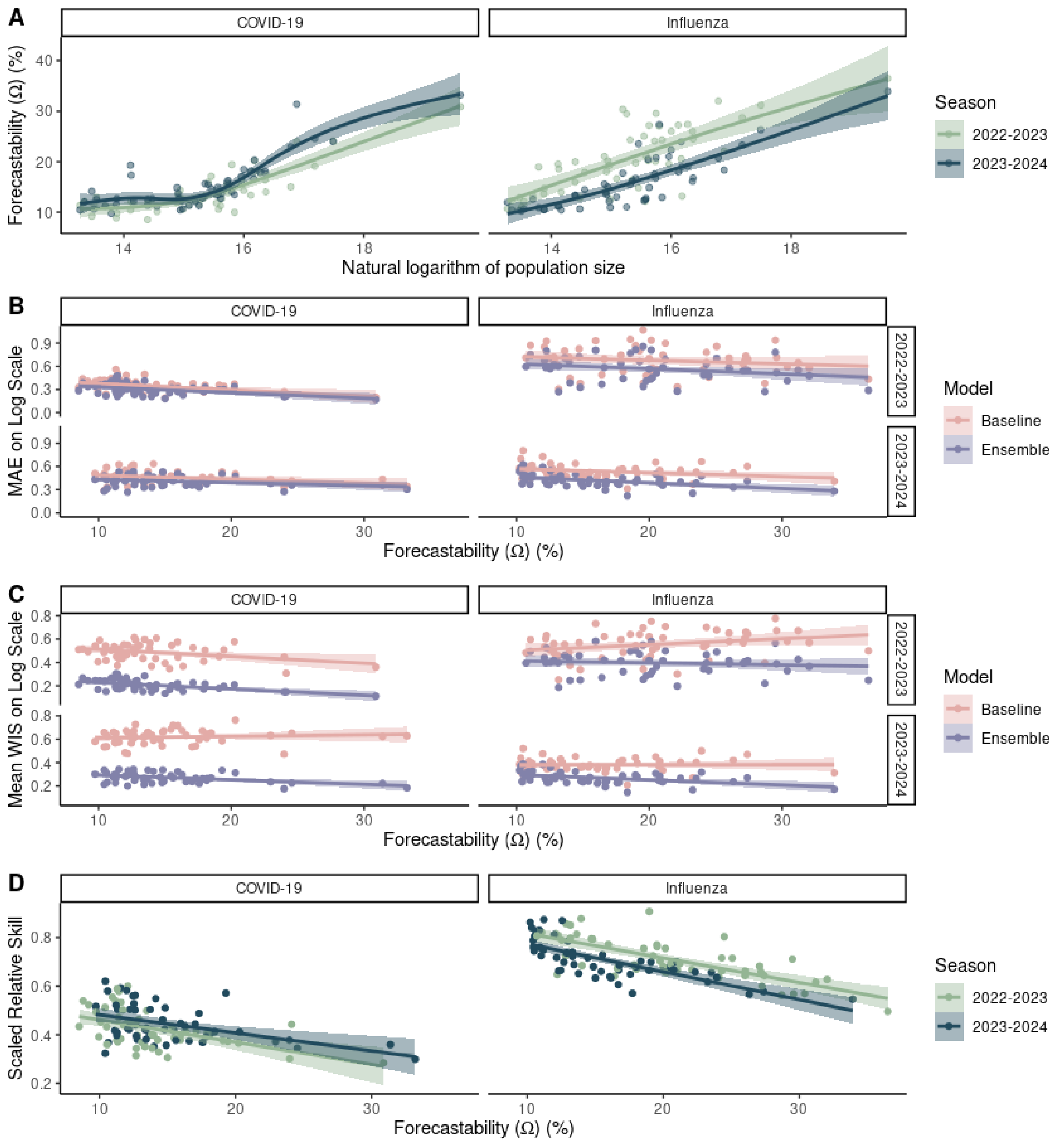


**Supplementary Figure 3.** The relationship between population size, forecastability, and forecast performance across two respiratory virus seasons for laboratory-confirmed (HHS/NHSN) COVID-19 and influenza admissions at the U.S. state and national scales. Here seasons are defined as July 1-June 30. **(A)** Forecastability (Ω) vs. the natural logarithm of population size. Forecast performance for the baseline and ensemble models as measured by: **(B)** MAE and **(C)** mean WIS and **(D)** scaled relative skill for the ensemble model.

**Supplementary Table 1.** Slope estimates from linear model fits for forecastability (Ω) vs. forecast performance for the ensemble and baseline models targeting laboratory-confirmed (HHS/NHSN) COVID-19 and influenza admissions at the U.S. state and national scales as shown in Supplementary Figure 3. Here seasons are defined as July 1-June 30. Values are shown with three significant figures. Bolded rows are those statistically significant at a $p = 0.05$ threshold.

| **Metric** | **Disease** | **Model** | **Season** | **Estimate (beta)** | **Standard error** | **Statistic** | **p-value** |
|---|---|---|---|---|---|---|---|
| MAE | COVID-19 | Baseline | **2022-2023** | **-0.00803** | **0.0025** | **-3.21** | **0.00231** |
| | | | **2023-2024** | **-0.00499** | **0.00229** | **-2.18** | **0.034** |
| | | Ensemble | **2022-2023** | **-0.00728** | **0.00215** | **-3.39** | **0.00138** |
| | | | **2023-2024** | **-0.00395** | **0.0019** | **-2.07** | **0.0432** |
| | Influenza | Baseline | 2022-2023 | -0.00451 | 0.00389 | -1.16 | 0.251 |
| | | | **2023-2024** | **-0.00498** | **0.00227** | **-2.2** | **0.0326** |
| | | Ensemble | **2022-2023** | **-0.00657** | **0.00326** | **-2.02** | **0.0491** |
| | | | **2023-2024** | **-0.00735** | **0.00192** | **-3.82** | **3.65e-4** |
| WIS | COVID-19 | Baseline | **2022-2023** | **-0.00586** | **0.00232** | **-2.53** | **0.0146** |
| | | | **2023-2024** | 0.00133 | 0.00178 | 0.75 | 0.457 |
| | | Ensemble | **2022-2023** | **-0.00551** | **0.00127** | **-4.35** | **6.74e-5** |
| | | | **2023-2024** | **-0.00395** | **0.00116** | **-3.42** | **0.00126** |
| | Influenza | Baseline | **2022-2023** | **0.00506** | **0.00248** | **2.04** | **0.0469** |
| | | | 2023-2024 | 8.14e-5 | 0.0015 | 0.0541 | 0.957 |
| | | Ensemble | 2022-2023 | -0.00179 | 0.0021 | -0.85 | 0.399 |
| | | | **2023-2024** | **-0.00442** | **0.00125** | **-3.53** | **9.16e-4** |
| Scaled relative skill | COVID-19 | Ensemble | **2022-2023** | **-0.00916** | **0.00222** | **-4.13** | **1.39e-4** |
| | | | **2023-2024** | **-0.00742** | **0.002** | **-3.71** | **5.27e-4** |
| | Influenza | | **2022-2023** | **-0.0101** | **0.00131** | **-7.71** | **4.71e-10** |
| | | | **2023-2024** | **-0.0115** | **0.00144** | **-7.97** | **1.83e-10** |
